# Expert consensus on barriers, facilitators and digital strategies to improve mental health service access for adolescents and young people living with HIV in Zambia 2025: A Delphi study

**DOI:** 10.64898/2026.01.23.26344682

**Authors:** Carlos Muleya, Jacqueline J. Folotiya, Peter J. Chipimo, Caitlin Baumhart, Cassidy Claassen, Naeem Dalal, Loyd Mulenga, Rosemary N. Likwa

**Author notes:** ***Corresponding author:*** Carlos Muleya.

## Abstract

**Background:** Adolescents and young people living with HIV (AYPLHIV) experience a high burden of depression, anxiety, behavioral difficulties, and psychosocial distress. These challenges undermine antiretroviral therapy (ART) adherence and long-term HIV outcomes. In Zambia, mental health services remain fragmented, centralized, and weakly integrated with HIV care. Expert consensus is needed to prioritize feasible system, service, and digital strategies to strengthen mental health access for AYPLHIV.

**Methods:** We conducted a two-round Delphi study with ten Zambian mental health experts (psychiatry, psychology, nursing, HIV program management, and digital health). Round One (September 1-10 2025) collected open-ended responses on barriers, facilitators, and mobile health (mHealth) strategies. Responses were thematically analyzed in Ligre 6.5.1 and synthesized into 29 candidate statements. In Round Two (September 15-20, 2025), experts rated each statement on a five-point Likert scale (1–5). Consensus was defined a priori as ≥70% rating a statement 4–5 and an interquartile range (IQR) ≤1.

**Results:** Consensus was reached for 27 of 29 statements (93%). Experts identified HIV- and mental-health–related stigma, poverty and transport costs, limited decentralization, low mental health literacy, harmful gender norms, and shortages of trained providers as key barriers to AYPLHIV accessing care. Priority facilitators included decentralizing services to primary and community levels, integrating mental health into ART clinics, strengthening youth-friendly and confidential counseling, expanding peer-support models, and improving multisector collaboration. mHealth solutions were strongly endorsed as confidential, scalable tools if they ensured robust data protection, offline or low-data functionality, multilingual and audio-visual content, and clear referral pathways to facility-based services.

**Conclusions:** This Delphi process generated clear expert consensus on pragmatic priorities for expanding youth mental health care within Zambia’s HIV program. The findings provide a framework for policymakers and implementers to integrate mental health and mHealth into HIV prevention, treatment, and care for AYPLHIV and to guide implementation and scale-up of youth-focused mental health interventions.

## Introduction

Adolescents and young people living with HIV (AYPLHIV) experience a high burden of common mental disorders, including depression, anxiety, and behavioral difficulties [1–3]. These conditions are associated with poorer antiretroviral therapy (ART) adherence, unsuppressed viral load, and increased risk of loss to follow-up, undermining long-term HIV outcomes [4–6]. In Zambia, adolescents living with HIV report substantial psychological distress, and symptoms of depression and anxiety are linked to suboptimal adherence to ART [5].

Despite this burden, mental health services in Zambia remain highly centralized and under-resourced, with few specialist providers, limited integration into HIV programs, and pronounced urban–rural inequities in access to care [7]. Recent policy developments—including the Mental Health Act of 2019 and the Zambia Digital Health Strategy 2022–2026—signal national commitment to strengthening rights-based, integrated mental health care and leveraging digital tools for service delivery [8,9]. However, implementation at district and community levels has been slow, and youth-focused mental health services remain scarce.

Digital and mobile health (mHealth) interventions are increasingly recognized as promising strategies to expand access to mental health support for young people in low- and middle-income countries, including through self-help tools, remote counseling, and task-sharing models [10–13]. Yet there is limited locally generated evidence on which strategies should be prioritized for AYPLHIV in Zambia and how they should be integrated with existing HIV service platforms.

Expert consensus can help prioritize contextually appropriate, feasible, and high-impact strategies for policy and program design. This Delphi study therefore aimed to: (1) identify expert consensus on key barriers and facilitators affecting AYPLHIV access to mental health services in Zambia; and (2) prioritize mHealth-supported strategies to strengthen youth mental health service delivery within HIV programs.

A separate qualitative manuscript (reported elsewhere) explores the lived experiences of AYPLHIV and policymakers; this Delphi paper focuses solely on expert perspectives and consensus.

## Methods Study design

We conducted a two-round Delphi study to obtain expert consensus on barriers, facilitators, and digital strategies to improve AYPLHIV mental health service access in Zambia. The Delphi rounds were conducted between September 1-20, 2025. Round One collected open-ended qualitative responses that were analyzed thematically; Round Two used structured Likert-scale ratings to quantify agreement. The conduct and reporting of the Round One qualitative component were informed by the Consolidated Criteria for Reporting Qualitative Research (COREQ) checklist [14].

### Participants and sampling

Participants were purposively sampled to include individuals with substantial experience in mental health, HIV service delivery, and/or digital health in Zambia. Eligibility criteria were: (i) at least five years of experience in clinical, programming, or policy roles relevant to mental health or HIV; (ii) current or recent involvement in adolescent or youth health programming; and (iii) willingness to participate in two iterative rounds.

We aimed to achieve diversity in professional background (psychiatry, psychology, mental health nursing, HIV program management, digital health) and institutional level (national, provincial, district, academic, and non-governmental). Ten experts were enrolled, including psychiatrists, a psychologist, mental health nurses, a provincial mental health coordinator, HIV program managers, and digital health implementers. Participants had 5–25 years of professional experience.

### Round One: qualitative item generation

In Round One, experts completed a structured, open-ended questionnaire administered electronically. They were asked to describe: (i) the main barriers preventing AYPLHIV from accessing mental health services; (ii) key facilitators or enabling factors to improve access and quality; and (iii) opportunities, risks, and design priorities for using mHealth and other digital tools to support AYPLHIV mental health.

Responses were imported into Ligre 6.5.1 (LIGRE, France) for thematic analysis. An inductive–deductive hybrid approach was used: initial open coding captured concepts emerging from the data, which were then grouped into categories informed by the study objectives (barriers, facilitators, digital strategies). Codes were iteratively refined, and related codes were clustered into themes and subthemes.

The study team then reformulated these themes into 29 concise, single-concept statements suitable for rating in Round Two. Statements were worded to maximize clarity, avoid double-barrelled phrasing, and reflect the core idea expressed by multiple experts.

### Round Two: consensus rating

In Round Two, the 29 statements were circulated to all ten experts using a self-administered online questionnaire. For each statement, experts were asked to rate their level of agreement on a five-point Likert scale, where 1 = strongly disagree, 2 = disagree, 3 = neutral, 4 = agree, and 5 = strongly agree.

For each item, we calculated the median, interquartile range (IQR), and the proportion of experts who rated the item as 4 or 5 (“agree” or “strongly agree”). Following published guidance on Delphi methodology [10–13], consensus was defined a priori as: (i) ≥70% of experts rating the statement as 4 or 5; and (ii) IQR ≤ 1, indicating low dispersion.

Statements meeting both criteria were considered to have reached consensus. Items that failed to meet either threshold were identified as non-consensus.

Because a high proportion of items met consensus criteria in Round Two and response distributions were stable, a third Delphi round was deemed unnecessary.

### Ethics

Ethical approval was obtained from the University of Zambia Biomedical Research Ethics Committee (UNZABREC; Reference 5864-2024) and the National Health Research Authority (NHRA; Reference NHRA-1679/04/11/2024). All participants provided written informed consent before participation. Data was anonymized for analysis and reporting.

## Results

The ten experts represented a range of professional roles and sectors, including clinical psychiatry, clinical psychology, mental health nursing, provincial mental health coordination, HIV program management, and digital health implementation. This mix of expertise ensured that both clinical and system-level dimensions of youth mental health were reflected in the consensus process.

Of the 29 statements rated in Round Two, 27 (93%) met the predefined consensus criteria of ≥70% agreement and IQR ≤ 1. Two statements did not reach consensus due to greater variability in ratings, indicating mixed views among experts. Statements clustered into four broad domains: (i) barriers to accessing mental health services; (ii) facilitators and system-strengthening opportunities; (iii) core features of mHealth and digital strategies; and (iv) cultural and multisector considerations. Table 1 below summarizes each statement with its median rating, IQR, percentage agreement, and consensus status.

**Table 1.**
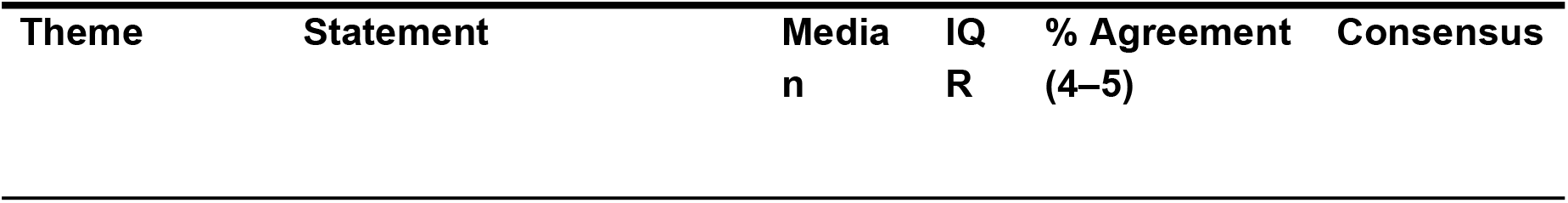
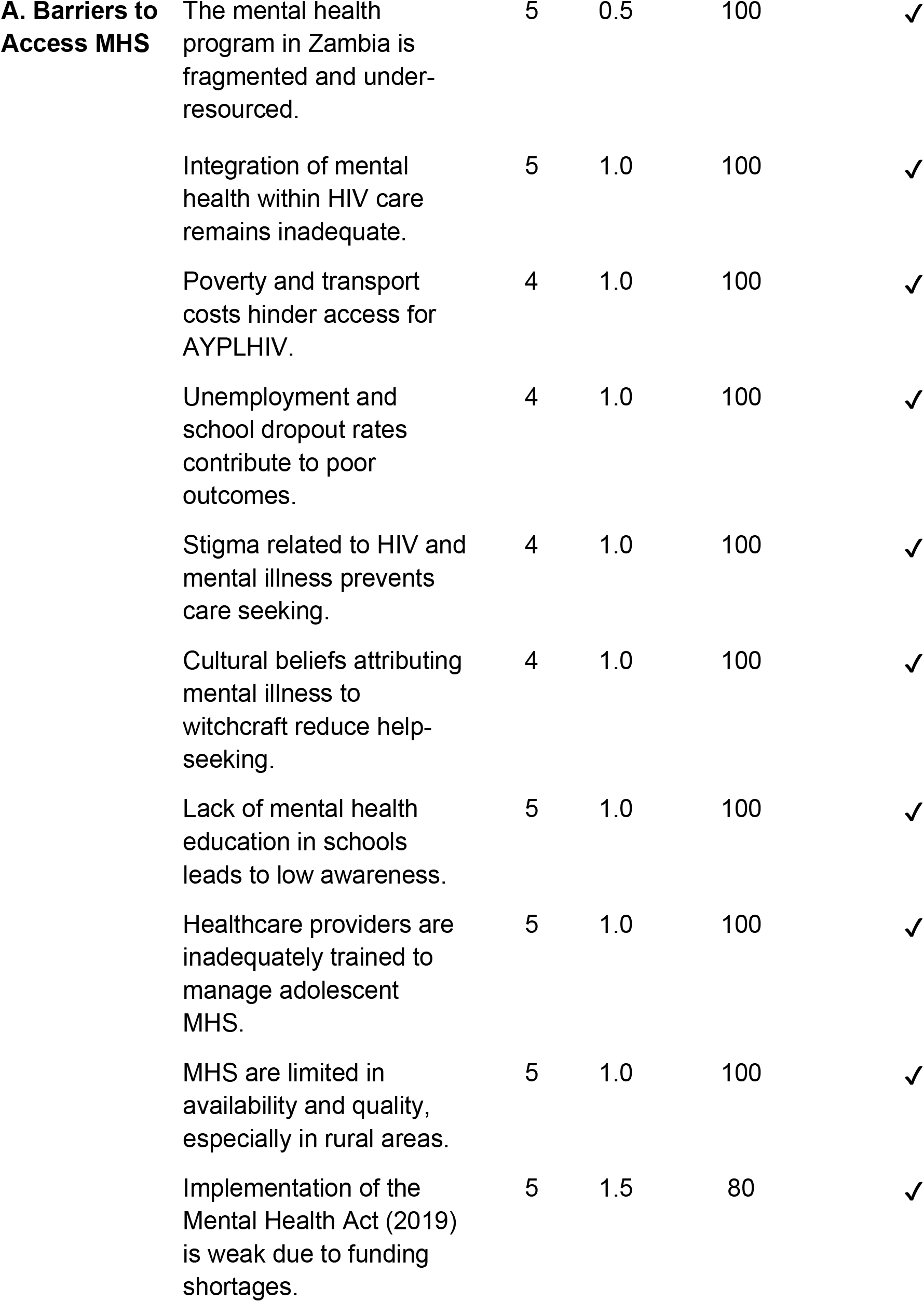
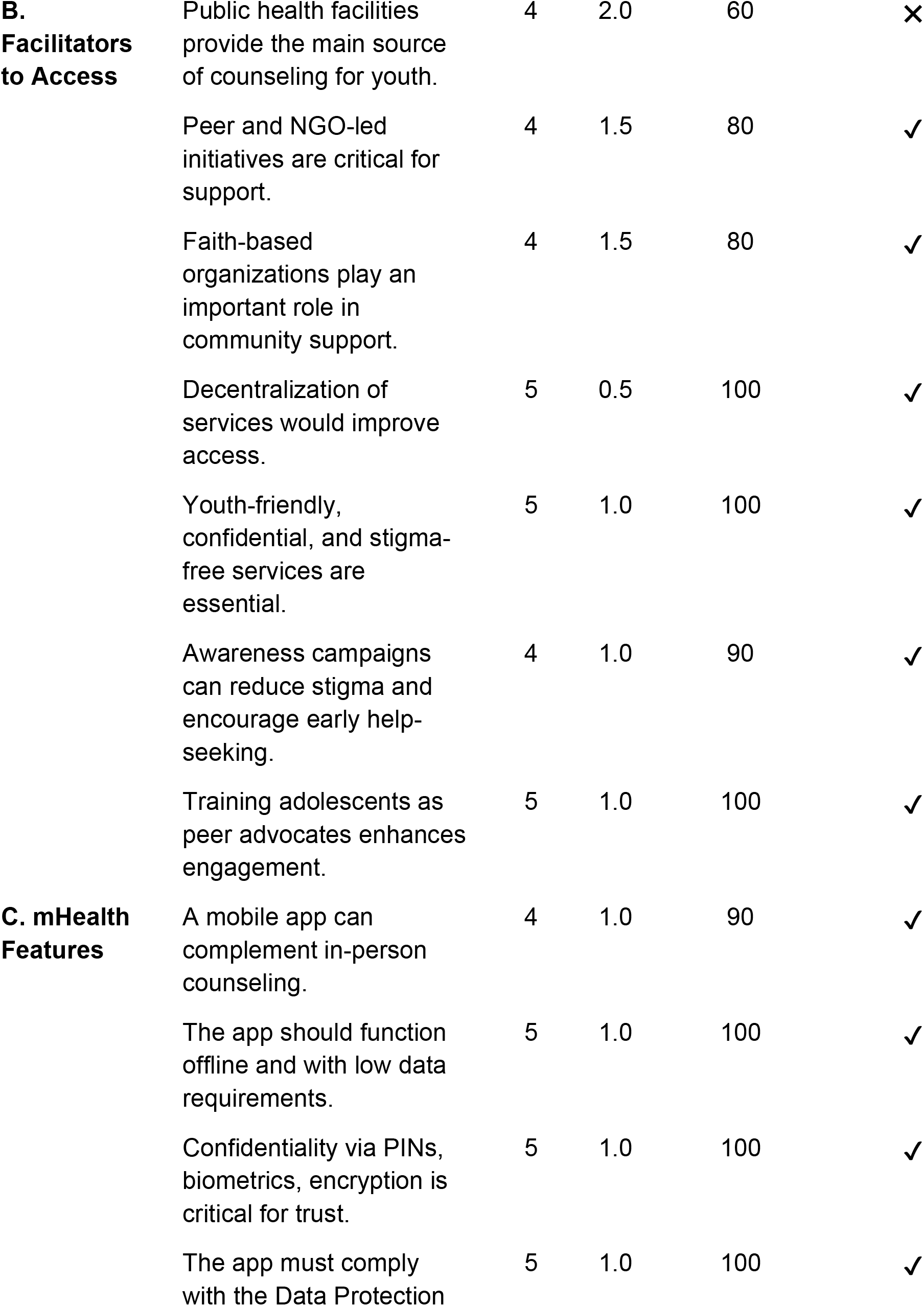
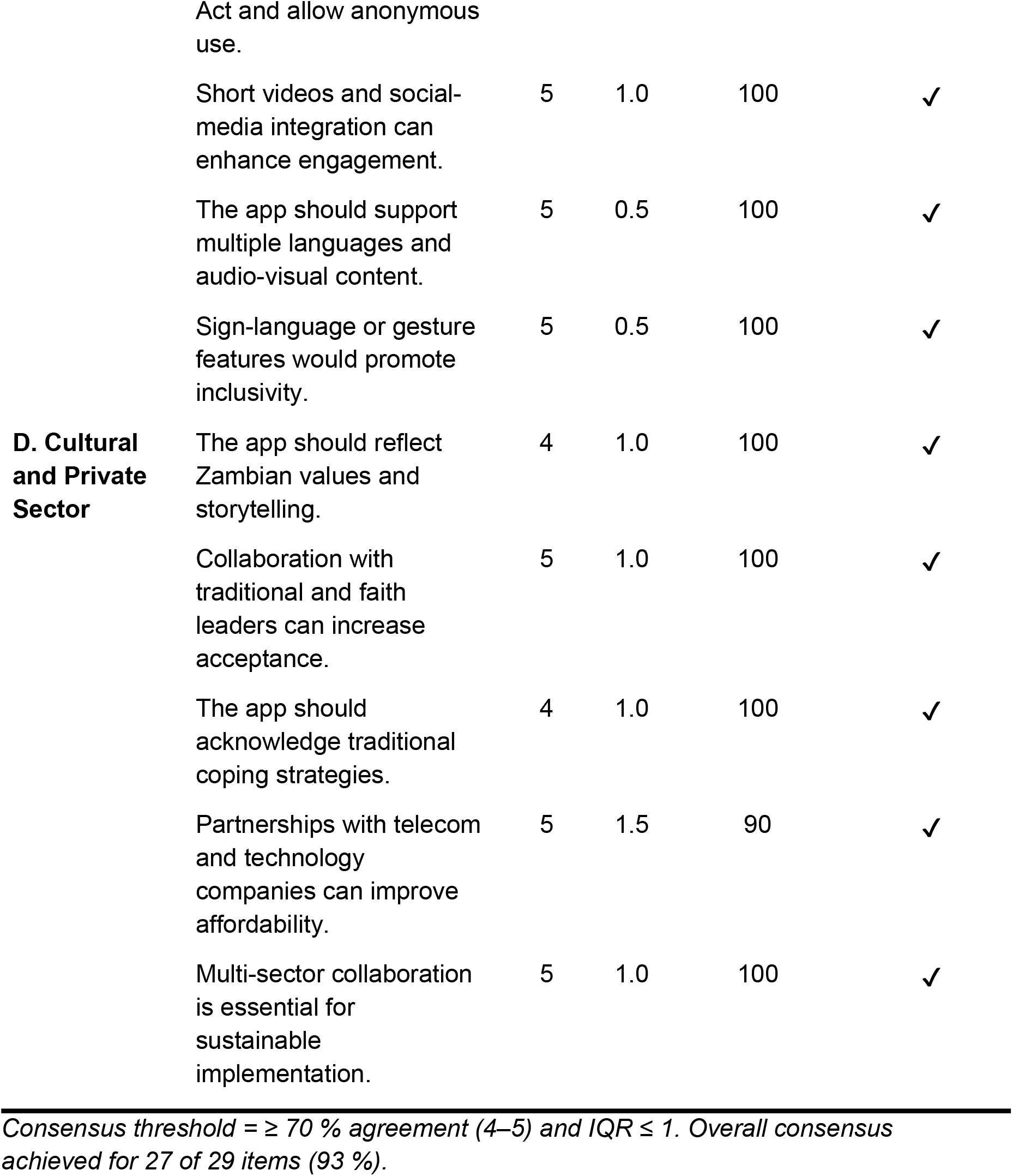
Summary of Delphi Round Two Consensus Results (n = 10)

### Barriers to mental health access

Experts strongly agreed that Zambia’s mental health system is fragmented and under-resourced, with services heavily concentrated in tertiary urban facilities and limited availability of youth-focused mental health care at primary and community levels. They identified stigma—both HIV-related and mental-health– related—as a central barrier, leading to “double stigma” that discourages AYPLHIV from disclosing distress or seeking help.

Socioeconomic barriers, particularly poverty and transport costs, were also highlighted as major constraints. Many AYPLHIV cannot afford regular travel to facilities, especially when mental health support requires multiple visits to centralized sites. Experts further noted low mental health literacy among adolescents, caregivers, and communities, which reinforces stigma and contributes to delayed or inappropriate help-seeking.

Cultural beliefs attributing mental illness to witchcraft, curses, or spiritual causes, and gender norms (e.g., that “men should not cry” or show emotional vulnerability), were recognized as important influences shaping help-seeking behavior. These beliefs can lead families to seek help from traditional or spiritual providers instead of—or before—formal health services.

Finally, experts emphasized shortages of trained mental health providers, particularly in rural districts, and weak operationalization of the Mental Health Act, due to insufficient funding and limited human resources, as structural barriers.

### Facilitators and enabling strategies

Consensus was reached on several key facilitators to improve AYPLHIV mental health access. Experts prioritized decentralization of services, including the integration of mental health assessment and basic interventions into primary health care and ART clinics. They agreed that bringing services closer to communities would reduce travel costs, increase continuity of care, and make it easier for AYPLHIV to receive both HIV and mental health support in a single setting.

Youth-friendly, confidential, and stigma-free services were consistently highlighted as essential. Experts stressed that adolescents need safe spaces and trusted providers with whom they can discuss sensitive issues without fear of judgment or breaches of confidentiality. Training adolescents and young adults as peer supporters or advocates was viewed as a promising strategy to normalize mental health conversations, reduce stigma, and enhance engagement.

Experts also recognized the important roles of peer-led initiatives, NGOs, and faith-based organizations in providing psychosocial support, especially in resource-limited settings. Public education and social and behavior change campaigns to improve mental health literacy and challenge stigma were seen as critical for creating an enabling environment.

Multisector collaboration—across health, education, social protection, youth, and telecommunications sectors—was identified as an important facilitator for sustainable mental health system strengthening.

### Digital and mHealth strategies

There was very strong consensus that mHealth solutions could play a critical complementary role in expanding mental health support for AYPLHIV. Experts agreed that mobile applications or SMS-based platforms could provide psychoeducation, coping skills, self-screening tools, and connection to human support (e.g., counselors, peer groups) in ways that are more accessible and less stigmatizing for young people.

Key design features endorsed by experts included: (i) confidentiality and data protection (end-to-end encryption, secure authentication, and adherence to the Data Protection Act); (ii) offline or low-data functionality, given connectivity constraints and the cost of mobile data; (iii) multilingual and inclusive content, including local languages and audio-visual formats; (iv) engaging formats such as short videos and interactive content; and (v) clear linkages to services, with triage and referral mechanisms for facility-based or emergency care.

Experts also endorsed exploring partnerships with telecommunications companies to reduce data costs and improve reach, and collaboration with community, traditional, and faith leaders to support acceptance and uptake of digital mental health tools.

### Non-consensus items

Two statements did not reach consensus. One related to whether counseling services currently offered in public health facilities are adequate; experts reported variable experiences across facilities and provinces, resulting in mixed ratings. The second concerned the strength of implementation of the Mental Health Act; while most experts felt implementation remained weak overall, there was disagreement about whether recent local initiatives represented meaningful progress.

These non-consensus items underline the heterogeneity of service availability and quality across Zambia and point to areas where more granular assessment is needed.

## Discussion

This Delphi study achieved strong expert consensus on key barriers, facilitators, and digital strategies for improving AYPLHIV mental health access in Zambia. The prioritized barriers—stigma, poverty, centralization, limited youth-friendly services, and workforce shortages—mirror evidence from sub-Saharan Africa showing that structural and social determinants strongly shape mental health outcomes and treatment engagement for adolescents living with HIV [1–5]. The emphasis on decentralization, task-sharing, and integration of mental health into HIV and primary care is consistent with the World Health Organization’s Mental Health Gap Action Programme (mhGAP) and global mental health frameworks that call for community-based, stepped-care models in low-resource settings [15,16].

Experts strongly endorsed digital and mHealth approaches as complementary tools to expand access to psychosocial support, provided that interventions are confidential, culturally appropriate, and aligned with existing health system workflows. This aligns with global evidence that digital technologies—including SMS, smartphone applications, and online platforms—can feasibly deliver mental health promotion, screening and low-intensity interventions, and support non-specialist providers in low- and middle-income countries [10–13]. At the same time, experts highlighted important constraints such as data costs, connectivity gaps, and language diversity, underscoring the need for offline-capable, low-bandwidth solutions co-designed with young users to ensure accessibility and acceptability.

The strong consensus around youth-friendly, confidential services and peer support resonates with studies demonstrating that stigma, fear of disclosure, and lack of trust in providers can deter AYPLHIV from seeking care, while peer-based interventions can improve engagement and adherence. [1–6] Our findings therefore reinforce the importance of combining structural and service-level reforms with social and behavioral interventions to address multi-level determinants of mental health.

The two non-consensus items point to substantial sub-national variation in service availability and quality. Rather than representing contradictions in expert views, they likely reflect differences in exposure to local initiatives and underscore the importance of district-level assessments and tailored implementation strategies.

A companion qualitative study with AYPLHIV and policymakers (reported elsewhere) identified overlapping themes related to stigma, structural barriers, and the potential of digital tools. Together, the Delphi and qualitative findings can inform the design of integrated intervention packages and implementation research efforts.

This study has several strengths. It used a structured Delphi methodology with clearly defined consensus thresholds, involved a diverse panel of experts from clinical, programmatic, and digital health backgrounds, and systematically transformed qualitative insights into quantifiable statements. The high proportion of items reaching consensus suggests that the identified priorities are widely endorsed among national experts.

Limitations include the relatively small number of experts (consistent with Delphi practice but inherently limited in representativeness) and the focus on expert perspectives rather than those of frontline providers or AYPLHIV themselves. While the consensus reflects expert judgment, implementation research will be required to test the feasibility, acceptability, and effectiveness of the proposed strategies. Additionally, we did not conduct a third Delphi round; however, given the high consensus rate and stable distributions, an additional round was unlikely to materially change the findings.

## Conclusions

This Delphi study identified strong expert consensus on the main barriers, facilitators, and digital strategies needed to strengthen mental health access for AYPLHIV in Zambia. Experts converged on the importance of addressing stigma, poverty, cultural beliefs, and workforce shortages; decentralizing and integrating mental health services within primary care and ART clinics; and developing secure, youth-centered mHealth tools that complement facility-based care.

These findings provide a pragmatic framework for policymakers and implementers seeking to integrate mental health into Zambia’s HIV response and to leverage digital technologies to close the mental health treatment gap for AYPLHIV.

## Recommendations

### Policy and system-level recommendations

- Decentralize mental health services by integrating screening and basic management into primary health care and ART clinics.
- Operationalize the Mental Health Act (2019) through clear implementation plans, budget allocations, and monitoring mechanisms.
- Expand and train the mental health workforce via task-sharing, using mhGAP-aligned training for nurses, clinical officers, lay counselors, and peer supporters [15].
- Strengthen multisector collaboration across health, education, social protection, youth, and telecommunications sectors to address social determinants and enable system-wide responses.

### Digital health recommendations

- Develop and test secure, youth-friendly mHealth platforms for AYPLHIV that provide psychoeducation, self-help tools, remote support, and clear referral pathways.
- Ensure robust confidentiality and data protection, low-data/offline functionality, and multilingual and inclusive design (including audio-visual and sign-language options) [10–13].
- Establish partnerships with telecommunications providers to reduce data costs and expand access.
- Integrate mHealth tools into routine HIV service delivery, including adolescent and youth-friendly services and community-based programs.

### Research recommendations

- Conduct implementation research to evaluate the acceptability, feasibility, and impact of decentralized and digital mental health interventions for AYPLHIV.
- Explore optimal models for integrating mental health into HIV service cascades at primary, secondary, and tertiary levels.
- Use participatory and co-design approaches to ensure that interventions and digital tools reflect AYPLHIV preferences and lived experiences.

## Data Availability

Full Round One qualitative responses and complete transcripts cannot be shared publicly because they contain detailed free?text information from a very small group of nationally identifiable mental?health experts in Zambia. Even after de?identification, there remains a high risk of deductive disclosure. The University of Zambia Biomedical Research Ethics Committee (UNZABREC Ref. 5864?2024) and the National Health Research Authority (NHRA Ref. NHRA?1679/04/11/2024) approved the study on the condition that these qualitative data would not be placed in an open public repository. Researchers who meet criteria for access to confidential data, as determined by UNZABREC and NHRA, may request access to de-identified qualitative data from the corresponding author (Carlos Muleya) for ethically approved secondary analyses.

## Acknowledgments

This work was supported by the Fogarty International Center of the National Institutes of Health (NIH) under Award Number 5D43TW012493 (ZENITH project). The content is solely the responsibility of the authors and does not necessarily represent the official views of the NIH, the Fogarty International Center, or the U.S. Department of Health and Human Services.

We thank all participating mental health experts for sharing their time and insights and acknowledge the broader ZENITH team and partners for their contributions to study design and coordination.

## Ethics statement

This study was approved by the University of Zambia Biomedical Research Ethics Committee (UNZABREC; Ref. 5864-2024) and the National Health Research Authority (NHRA; Ref. NHRA-1679/04/11/2024). All participants provided written informed consent. The study adhered to relevant national and institutional ethical guidelines.

## Data availability

De-identified Delphi data (Round Two ratings) are available from the corresponding author upon reasonable request, subject to ethical and privacy considerations.

## Author contributions

CM conceived and designed the study, coordinated data collection, conducted analysis, and drafted the manuscript. JF, JB, PJC, BMK, LM, PS, CN, CB, CC, ND, LM and RL contributed to study design refinement, interpretation of findings, and critical revision of the manuscript. All authors read and approved of the final manuscript.

## References

1. Vreeman RC, McCoy BM, Lee S. Mental health challenges among adolescents living with HIV. J Int AIDS Soc. 2017;20(Suppl 3):21497.

2. Mellins CA, Malee KM. Understanding the mental health of youth living with perinatal HIV infection: lessons learned and current challenges. J Int AIDS Soc. 2013;16(1):18593.

3. Dessauvagie AS, Jörns-Presentati A, Napp A-K, Stein DJ, Jonker D, Breet E, et al. The prevalence of mental health problems in sub-Saharan adolescents living with HIV: a systematic review. Glob Ment Health. 2020;7:e18.

4. Olashore AA, Paruk S, Akanni OO, Tomita A, Chiliza B. Psychiatric disorders in adolescents living with HIV and association with antiretroviral therapy adherence in sub-Saharan Africa: a systematic review and meta-analysis. AIDS Behav. 2021;25(6):1711–28.

5. Okawa S, Yasuoka J, Ishikawa N, Poudel KC, Ryu S, El-sahn M, et al. Psychological well-being and adherence to antiretroviral therapy among adolescents living with HIV in Zambia. AIDS Care. 2018;30(6):734–42.

6. UNICEF Eastern and Southern Africa. Mental Health and Antiretroviral Treatment Adherence among Adolescents Living with HIV: Evidence on Risk Pathways and Protective Factors. Nairobi: UNICEF; 2021.

7. World Health Organization. Mental Health Atlas 2020: Zambia Country Profile. Geneva: World Health Organization; 2021.

8. Republic of Zambia. Mental Health Act, 2019 (Act No. 6 of 2019). Lusaka: Government of the Republic of Zambia; 2019.

9. Ministry of Health (Zambia). Zambia Digital Health Strategy 2022–2026. Lusaka: Government of the Republic of Zambia; 2022.

10. Naslund JA, Aschbrenner KA, Araya R, Marsch LA, Unützer J, Patel V, et al. Digital technology for treating and preventing mental disorders in low-income and middle-income countries: a narrative review of the literature. Lancet Psychiatry. 2017;4(6):486–500.

11. Donker T, Petrie K, Proudfoot J, Clarke J, Birch MR, Christensen H. Smartphones for smarter delivery of mental health programs: a systematic review. J Med Internet Res. 2013;15(11):e247.

12. Grist R, Porter J, Stallard P. Mental health mobile apps for preadolescents and adolescents: a systematic review. J Med Internet Res. 2017;19(5):e176.

13. Kola L, Abiona D, Adefolarin AO, Ben-Zeev D. Mobile phone use and acceptability for the delivery of mental health information among perinatal adolescents in Nigeria: survey study. JMIR Ment Health. 2021;8(1):e20314.

14. Tong A, Sainsbury P, Craig J. Consolidated criteria for reporting qualitative research (COREQ): a 32-item checklist for interviews and focus groups. Int J Qual Health Care. 2007;19(6):349–57.

15. World Health Organization. mhGAP Intervention Guide for Mental, Neurological and Substance Use Disorders in Non-specialized Health Settings, Version 2.0. Geneva: World Health Organization; 2016.

16. Patel V, Saxena S, Lund C, Thornicroft G, Baingana F, Bolton P, et al. The Lancet Commission on global mental health and sustainable development. Lancet. 2018;392(10157):1553–98.

